## Appendix Tables for "Social support and ideal cardiovascular health in urban Jamaica: a cross-sectional study"

### Appendix with Supplementary Tables

---

**Authors:**

Alphanso L. Blake[1, 4], Nadia R. Bennett[1], Joette A. McKenzie[1], Marshall K. Tulloch-Reid[1], Ishtar Govia[1], Shelly R. McFarlane[1], Renee Walters[1], Damian K. Francis[2], Rainford J. Wilks[1], David R. Williams[3], Novie O. Younger-Coleman[1*], Trevor S. Ferguson[1*]

**INSTITUTIONS**

[1]Caribbean Institute for Health Research, The University of the West Indies, Mona, Kingston, Jamaica

[2]School of Health and Human Performance, Georgia College and State University, Milledgeville, GA, USA

[3]Department of Social and Behavioral Sciences, Harvard T. H. Chan School of Public Health, Boston, MA, USA

[4]School of Clinical Medicine and Research, The Faculty of Medical Sciences, The University of the West Indies, Mona, Nassau, The Bahamas

***Corresponding author:***

Trevor Ferguson

*These authors are joint senior authors on this manuscript.: TSF, NYC

**Table S1: Mean values for participant characteristics by sex (estimates weighted for survey design; no imputation)**

| Characteristic | Male n= 279 | Female n= 562 | Total N=841 | p-value for sex difference |
|---|---|---|---|---|
| *Mean for Continuous Variables* | Mean ± SE | Mean ± SE | Mean ± SE | |
| **Age (years)** | 38.5 ± 0.1 | 38.1 ± 0.9 | 38.3±0.1 | 0.015 |
| **Height (cm)** | 173.4 ± 0.6 | 162.8 ± 0.6 | 167.9±0.5 | <0.001 |
| **Weight (kg)** | 77.4 ± 1.1 | 78.8 ± 1.2 | 78.1±0.8 | 0.383 |
| **Body Mass Index (kg/m2)** | 25.6 ±0.4 | 29.6 ± 0.4 | 27.7±0.3 | <0.001 |
| **Systolic Blood Pressure (mmHg)** | 127.6 ±0.8 | 121.7 ±1.4 | 124.5±0.9 | <0.001 |
| **Diastolic Blood Pressure (mmHg)** | 81.6 ±0.6 | 80.8 ±0.9 | 81.2±0.6 | 0.399 |
| **Fasting Glucose (mmol/L)** | 5.8 ± 0.4 | 5.5 ±0.1 | 5.6±0.2 | 0.342 |
| **Total Cholesterol (mmol/L)** | 4.2 ± 0.1 | 4.2 ± 0.1 | 4.2±0.1 | 0.854 |
| **ICH Score** | 3.7 ± 0.1 | 3.5 ± 0.1 | 3.6 ± 0.1 | 0.156 |
| **Social Support Score (PCA)** | 0.22 ± 0.1 | -0.15 ± 0.1 | 0.0 ± 0.1 | 0.004 |
| **Categorical Variables** | **% ± SE** | **% ± SE** | **% ± SE** | |
| **Education** | | | | 0.851 |
| Less than High School | 12.4±1.2 | 11.2±1.6 | 11.8±1.1 | |
| High School | 53.5±3.6 | 53.3±3.7 | 53.4±3.1 | |
| More than High School | 34.1±4.0 | 35.5±4.1 | 34.8±3.4 | |
| **Median Land Value** | | | | 0.264 |
| Lower Tertile | 46.4±7.2 | 50.6±7.9 | 48.6±7.4 | |
| Middle Tertile | 38.7±6.5 | 34.4±6.3 | 36.5±6.2 | |
| Upper Tertile | 14.9±4.8 | 15.1±5.3 | 15.0±4.9 | |
| **Community Poverty** | | | | 0.260 |
| Lower Tertile | 46.1±8.1 | 40.4±9.0 | 43.1±8.4 | |
| Middle Tertile | 21.4±6.0 | 22.0±6.4 | 21.7±6.1 | |
| Lower Tertile | 32.5±5.6 | 37.5±7.6 | 35.1±6.5 | |

*ICH – Ideal Cardiovascular Health; PCA- Principal Component Analysis
ICH Score = sum of number of individual ICH components
Social Support Score = PCA derived score from number of friends, number of friends willing to give advice and number of willing to provide loans.
Median Land Value – median property value for community based on data from National Land Agency
Community poverty – based on data from Jamaica Poverty May 2019

**Table S2A: Population age-sex numbers and proportions for urban Jamaica**

| Age | Population total - Urban Males<br><br>n | Population total - Urban Females<br>n | Percentage of Males in age groups<br>% | Percentage of Females in age groups<br>% | Percentage of Total in age groups<br>% |
|---|---|---|---|---|---|
| 15-24 | 140865 | 143549 | 27.2 | 25.0 | 26.0 |
| 25-34 | 111432 | 124757 | 21.5 | 21.7 | 21.6 |
| 35-44 | 93785 | 106845 | 18.1 | 18.6 | 18.4 |
| 45-54 | 75726 | 86588 | 14.6 | 15.1 | 14.9 |
| 55-64 | 48569 | 52389 | 9.4 | 9.1 | 9.2 |
| 65-74 | 27476 | 31060 | 5.3 | 5.4 | 5.4 |
| 75+ | 19703 | 29171 | 3.8 | 5.1 | 4.5 |
| Total | 517556 | 574359 | 100 | 100 | 100 |

**Table S2B: Sample age-sex numbers and proportions, weighted age-sex numbers, and proportions**

| **Age Groups** | **Sample total - Urban Males**<br><br>**n** | **Sample total - Urban Females**<br><br>**n** | **Percentage of Males in sample age groups**<br><br>**%** | **Percentage of Females in sample age groups**<br><br>**%** | **Percentage of Total in sample age groups**<br><br>**%** | **Weighted percentages of Urban Males in age groups**<br><br>**%** | **Weighted percentages of Urban Females in age groups**<br><br>**%** | **Weighted percentages of Urban residents in age groups**<br>**%** |
|---|---|---|---|---|---|---|---|---|
| **15-24** | 50 | 68 | 17.6 | 12.1 | 13.9 | 27.2 | 25.7 | 26.4 |
| **25-34** | 43 | 82 | 15.4 | 14.6 | 14.9 | 21.2 | 22.2 | 21.7 |
| **35-44** | 36 | 92 | 12.9 | 16.0 | 15.0 | 18.1 | 19.2 | 18.6 |
| **45-54** | 52 | 109 | 18.6 | 19.2 | 19.0 | 15.0 | 15.9 | 15.4 |
| **55-64** | 39 | 97 | 13.6 | 17.3 | 16.1 | 9.5 | 9.2 | 9.3 |
| **65-74** | 35 | 77 | 12.5 | 13.5 | 13.2 | 5.3 | 5.4 | 5.3 |
| **75+** | 27 | 42 | 9.3 | 7.3 | 8.0 | 3.7 | 2.6 | 3.1 |
| **Total** | 282 | 567 | 100 | 100 | 100 | 100 | 100 | 100 |

**Table S3: Prevalence of ICH-5 by Social Support Score Tertiles and by Socioeconomic Status (SES) Categories**

| SES Variable | Low | Middle | High | P-value for difference in proportions |
|---|---|---|---|---|
| **Males and Females** | | | | |
| Social Support Score[1] | 33.0 | 22.6 | 23.7 | 0.121 |
| Education Level[2] | 16.0 | 29.1 | 26.5 | 0.046 |
| Median Land Value | 30.9 | 19.6 | 29.9 | 0.020 |
| Poverty[3] | 22.3 | 23.1 | 34.2 | 0.056 |
| **Males** | | | | |
| Social Support Score | 44.3 | 10.4 | 20.8 | <0.001 |
| Education Level | 15.8 | 31.8 | 19.7 | 0.004 |
| Median Land Value | 31.4 | 17.3 | 29.7 | 0.041 |
| Poverty | 20.6 | 23.5 | 34.2 | 0.065 |
| **Females** | | | | |
| Social Support Score | 23.8 | 32.7 | 26.9 | 0.448 |
| Education Level | 16.2 | 26.6 | 32.5 | 0.087 |
| Median Land Value | 30.5 | 22.0 | 30.1 | 0.157 |
| Poverty | 24.0 | 22.6 | 34.2 | 0.177 |

[1]Social support score, median land value and poverty are categorized into tertiles.
[2]Education is categorized as less the high school, high school and more than high school.
[3]Poverty is categorized as proportion of persons experiencing poverty in a community so that higher categories indicate lower socioeconomic status.

**Table S4: Mean Social Support Score (from PCA) by Socioeconomic Status (SES) Categories**

| SES Variable | Low Mean | Middle Mean | High Mean | P-value for difference in means |
|---|---|---|---|---|
| Education Level | -0.18 | -0.16 | 0.43 | <0.001 |
| Median Land Value | -0.13 | 0.28 | 0.00 | <0.001 |
| Poverty | 0.03 | 0.39 | -0.17 | 0.016 |

**Table S5: Prevalence of Ideal Cardiovascular Health (ICH) Characteristics by Socioeconomic Status (SES) Level**

| ICH Characteristic | SES Category | | | P-value for difference in means |
|---|---|---|---|---|
| **Education** | **Less than High School** | **High School** | **More than High School** | |
| Normal BMI | 31 (22 – 40) | 40 (33 – 46) | 31 (23 – 39) | 0.286 |
| Non-smoker | 83 (73 – 92) | 77 (72 – 82) | 87 (81 – 94) | 0.076 |
| Normal glucose | 45 (35 – 54) | 68 (62 – 74) | 74 (64 – 86) | <0.001 |
| Normal blood pressure | 14 (6 – 22) | 38 (32 – 44) | 36 (27 – 45) | 0.004 |
| Adequate physical activity | 45 (35 – 54) | 40 (34 – 46) | 41 (33 – 49) | 0.652 |
| Healthy diet | 24 (17 – 31) | 19 (14 – 24) | 17 (12 – 22) | 0.248 |
| Normal Cholesterol | 64 (53 – 74) | 83 (79 – 88) | 76 (68 – 84) | 0.005 |
| **Median Land Value** | **Lower Tertile** | **Middle Tertile** | **Upper Tertile** | |
| Normal BMI | 38 (33 – 44) | 29 (23 – 35) | 43 (36 – 50) | 0.021 |
| Non-smoker | 77 (72 – 83) | 86 (79 – 93) | 82 (74 – 90) | 0.230 |
| Normal glucose | 71 (66 – 77) | 64 (51 – 77) | 64 (56 – 72) | 0.260 |
| Normal blood pressure | 34 (29 – 40) | 35 (25 – 45) | 33 (15 – 51) | 0.969 |
| Adequate physical activity | 40 (33 – 47) | 46 (39 – 53) | 29 (21 – 37) | 0.012 |
| Healthy diet | 16 (11 – 20) | 21 (15 – 27) | 26 (16 – 35) | 0.157 |
| Normal cholesterol | 87 (84 – 90) | 69 (62 – 76) | 74 (63 – 85) | <0.001 |
| **Community Poverty** | **Lower Tertile** | **Middle Tertile** | **Upper Tertile** | |
| Normal BMI | 35 (30 – 40) | 30 (24 – 36) | 40 (33 – 47) | 0.060 |
| Non-smoker | 82 (78 – 86) | 88 (79 – 97) | 76 (69 – 83) | 0.156 |
| Normal glucose | 61 (51 – 71) | 70 (55 – 84) | 74 (70 – 79) | 0.039 |
| Normal blood pressure | 32 (23 – 41) | 39 (29 – 49) | 34 (28 – 41) | 0.591 |
| Adequate physical activity | 39 (31 – 47) | 46 (38 – 53) | 39 (32 – 47) | 0.450 |
| Healthy diet | 25 (18 – 31) | 13 (8 – 19) | 16 (10 – 21) | 0.048 |
| Normal cholesterol | 74 (68 – 81) | 71 (59 – 82) | 88 (85 – 92) | <0.001 |

**Table S6: Prevalence of Ideal Cardiovascular Health Characteristics ICH Characteristics by Social Support Tertile**

| ICH Characteristic | Tertile 1 % (CI) | Tertile 2 % (CI) | Tertile 3 % (CI) | P-value (difference between groups) |
|---|---|---|---|---|
| **Males and Females** | | | | |
| Normal BMI | 37.4 (29.8 – 45.1) | 38.9 (30.9 – 46.8) | 32.8 (25.4 – 40.2) | 0.491 |
| Non-Smoker | 78.8 (72.7 – 84.8) | 80.3 (74.3 – 86.3) | 83.2 (76.8 – 89.7) | 0.616 |
| Normal Glucose | 68.7 (60.2 – 77.2) | 63.9 (54.9 – 72.9) | 68.9 (58.8 – 78.9) | 0.647 |
| Normal Blood Pressure | 38.5 (31.3 – 45.7) | 33.1 (26.2 – 40.0) | 31.5 (22.4 – 40.7) | 0.273 |
| Adequate Physical Activity | 40.1 (30.9 – 49.0) | 39.3 (32.4 – 46.2) | 41.3 (33.7 – 48.9) | 0.932 |
| Healthy Diet | 15.9 (10.1 – 21.8) | 22.8 (16.4 – 29.3) | 19.7 (15.2 – 24.1) | 0.144 |
| Normal Cholesterol | 83.8 (77.6 – 90.1) | 79.9 (72.6 – 87.2) | 73.8 (67.3 – 80.3) | 0.112 |
| **Males** | | | | |
| Normal BMI | 53.3 (40.7 – 65.9) | 44.1 (33.3 – 55.1) | 39.4 (25.7 – 53.0) | 0.414 |
| Non-Smoker | 76.7 (64.7 – 88.8) | 60.9 (49.5 – 72.2) | 74.3 (64.9 – 83.8) | 0.111 |
| Normal Glucose | 72.3 (61.7 – 83.1) | 59.2 (45.9 – 72.6) | 67.1 (54.7 – 79.2) | 0.385 |
| Normal Blood Pressure | 37.9 (27.9 – 47.9) | 22.7 (8.4 – 37.1) | 26.6 (17.4 – 35.7) | 0.136 |
| Adequate Physical Activity | 58.9 (46.4 – 71.5) | 48.7 (33.4 – 64.1) | 48.6 (38.3 – 58.9) | 0.416 |
| Healthy Diet | 17.1 (7.9 – 26.1) | 18.8 (8.1 – 29.5) | 14.4 (8.6 – 20.2) | 0.759 |
| Normal Cholesterol | 80.0 (69.8 – 90.2) | 81.1 (71.1 – 91.2) | 77.7 (67.4 – 87.9) | 0.887 |
| **Females** | | | | |
| Normal BMI | 24.7 (16.7 – 32.7) | 34.5 (25.4 – 43.5) | 25.6 (19.8 – 31.5) | 0.298 |
| Non-Smoker | 80.4 (73.9 – 86.9) | 96.4 (93.4 – 99.3) | 92.8 (88.1 – 97.7) | <0.001 |
| Normal Glucose | 65.8 (55.5 – 76.0) | 67.8 (55.4 – 80.2) | 70.9 (60.6 – 81.2) | 0.784 |
| Normal Blood Pressure | 40.1(29.1 – 49.0) | 41.7 (32.9 – 50.5) | 40.1 (23.9 – 50.0) | 0.842 |
| Adequate Physical Activity | 24.7 (16.4 – 33.1) | 31.6 (20.8 – 42.3) | 33.4 (25.7 – 41.2) | 0.206 |
| Healthy Diet | 15.1 (7.6 – 22.4) | 26.2 (17.5 – 34.9) | 25.4 (18.9 – 31.8) | 0.136 |
| Normal Cholesterol | 86.8 (79.9 – 93.6) | 78.8 (69.6 – 88.0) | 69.6 (62.9 – 76.2) | 0.015 |

**Table S7: Odds ratio for unit change in social support score for each ideal cardiovascular Health Characteristics in bivariate models.**

| ICH Characteristic | Males and Females OR (95% CI) | Males OR (95% CI) | Females OR (95% CI) |
|---|---|---|---|
| Normal BMI | 1.0 (0.87 – 1.14) | 0.93 (0.76 – 1.13) | 1.04 (0.94 – 1.16) |
| Non-smoker | 0.91 (0.76 – 1.09) | 0.87 (0.69 – 1.06) | 1.94 (1.25 – 3.01) ** |
| Normal glucose | 0.97 (0.82 – 1.14) | 0.94 (0.77 – 1.14) | 1.06 (0.85 – 1.32) |
| Normal blood pressure | 0.97 (0.84 – 1.15) | 0.96 (0.77 – 1.21) | 1.07 (0.91 – 1.26) |
| Adequate physical activity | 1.0 (0.87 – 1.15) | 0.89 (0.75 – 1.06) | 1.10 (0.90 – 1.35) |
| Healthy diet | 1.05 (0.92 – 1.20) | 0.93 (0.75 – 1.16) | 1.26 (1.07 – 1.48) ** |
| Normal cholesterol | 0.90 (0.78 – 1.04) | 1.0 (0.82 – 1.23) | 0.74 (0.62 – 0.90) ** |

*p<0.05; **p<0.01; ***p<0.001

**Table S8: Odds ratio for unit change in social support score for each ideal cardiovascular Health Characteristics in multivariable models[1].**

| ICH Characteristic | Males<br>OR (95% CI) | Females<br>OR (95% CI) |
|---|---|---|
| Normal BMI | 0.96 (0.81 – 1.14) | 1.18 (1.02 – 1.34) * |
| Non-smoker | 0.80 (0.66 – 0.99) * | 1.73 (1.32 – 2.66) * |
| Normal glucose | 0.88 (0.69 – 1.11) | 1.16 (0.91 – 1.47) |
| Normal blood pressure | 0.98 (0.80 – 1.20) | 1.20 (0.97 – 1.50) |
| Adequate physical activity | 0.91 (0.76 – 1.08) | 1.07 (0.80 – 1.44) |
| Healthy diet | 0.90 (0.69 – 1.17) | 1.20 (1.00 – 1.44) * |
| Normal cholesterol | 1.04 (0.81 – 1.33) | 0.82 (0.69 – 0.98) * |

*p<0.05; **p<0.01; ***p<0.001

[1]Adjusted for age, education level, median property value and community poverty.

**Table S9: Survey weighted multivariable model without imputations[1] showing the odds of having five or more ideal cardiovascular health characteristics (ICH-5) for males and females with their corresponding 95% CI and p-value.**

| Variables | Females<br>Odds ratio (95% CI) | P-value | Males<br>Odds ratio (95% CI) | P-value |
|---|---|---|---|---|
| Social Support Score | 1.3 (1.1 – 1.6) | 0.004 | 0.7 (0.5 – 0.9) | 0.003 |
| Age | 0.9 (0.9-1.0) | 0.001 | 1.0 (0.9 – 1.0) | 0.005 |
| Education Category | | | | |
| Less than High School | Reference | Reference | Reference | Reference |
| High School | 0.4 (0.1 – 1.6) | 0.185 | 1.3 (0.6 – 3.1) | 0.486 |
| More Than High School | 0.7 (0.2 – 2.0) | 0.485 | 0.7 (0.2 – 2.4) | 0.555 |
| Land Value Category | | | | |
| Lower Category | Reference | Reference | Reference | Reference |
| Middle category | 1.3 (0.6 – 2.6) | 0.528 | 0.4 (0.1 – 1.3) | 0.130 |
| Upper Category | 2.5 (1.1 – 5.4) | 0.026 | 2.5 (0.8 – 7.5) | 0.096 |
| Community Poverty Category | | | | |
| Lower Category | Reference | Reference | Reference | Reference |
| Middle Category | 1.2 (0.5 – 2.8) | 0.705 | 5.2 (1.5 – 17.9) | 0.010 |
| Upper Category | 2.1 (0.7 – 6.0) | 0.168 | 1.9 (1.0 – 3.7) | 0.055 |

[1]Separate models created for 245 males and 495 females with ICH-5 as outcome, social support score and main exposure variable and adjusting for age, education, median land value and community poverty.

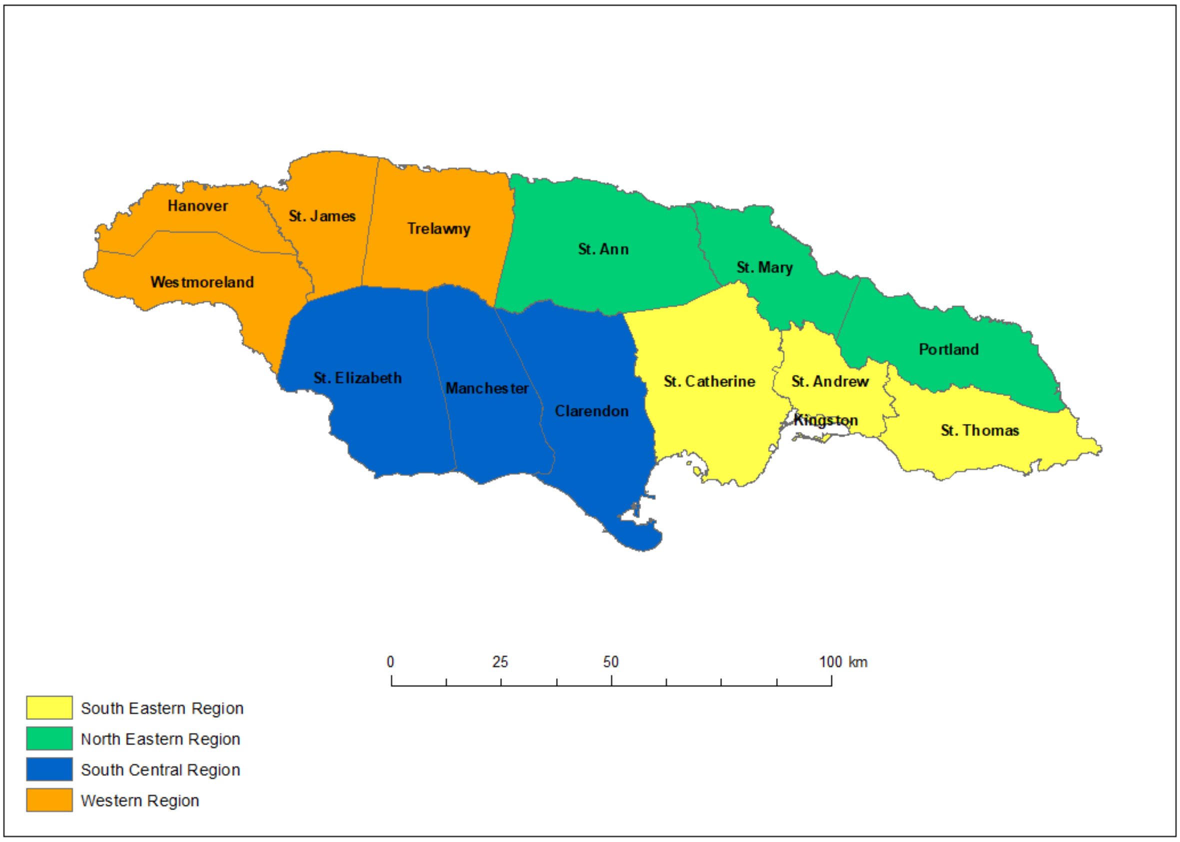

Figure S1: Map of Jamaica showing parishes and health regions.
The study was conducted in communities selected from Jamaica's Southeast Health Region (yellow-coloured parishes on the map)
